# Impacts of the COVID-19 Pandemic on Cardiac Rehabilitation Delivery around the World

**DOI:** 10.1101/2020.11.11.20230045

**Authors:** Gabriela Lima de Melo Ghisi, Zhiming Xu, Xia Liu, Ana Mola, Robyn Gallagher, Abraham Samuel Babu, Colin Yeung, Susan Marzolini, John Buckley, Paul Oh, Aashish Contractor, Sherry L Grace

## Abstract

**Background:** To investigate impacts of COVID-19 on CR delivery around the globe, including effects on providers and patients.

**Methods:** In this cross-sectional study, a piloted survey was administered to CR programs globally via REDCap from April-June/2020. The 50 members of the ICCPR and personal contacts facilitated program identification.

**Results:** Overall, 1062(18.3% program response rate) responses were received from 70/111(63.1% country response rate) countries in the world with existent CR programs. Of these, 367(49.1%) programs reported they had stopped CR delivery, and 203(27.1%) stopped temporarily (mean=8.3±2.8weeks). Alternative models were delivered in 322(39.7%) programs, primarily through low-tech modes (n=226,19.3%). 353(30.2%) respondents were re-deployed, and 276 (37.3%) felt the need to work due to fear of losing their job, despite the perceived risk of contracting COVID-19 (mean=30.0%±27.4/100). 266(22.5%) reported anxiety, 241(20.4%) were concerned about exposing their family, 113(9.7%) reported increased workload to transition to remote delivery, and 105(9.0%) were juggling caregiving responsibilities during business hours. Patients were often contacting staff regarding grocery shopping for heart-healthy foods (n=333,28.4%), how to use technology to interact with the program (n=329,27.9%), having to stop their exercise because they have no place to exercise (n=303,25.7%), and their risk of death from COVID-19 due to pre-existing cardiovascular disease (n=249,21.2%). Respondents perceived staff (n=488,41.3%) and patient (n=453,38.6%) personal protective equipment, as well as COVID-19 screening (n=414,35.2%) and testing (n=411,35.0%) as paramount to in-person service resumption.

**Conclusion:** Approximately 4400 programs ceased service delivery. Those that remain open are implementing new technologies to ensure their patients receive CR safely, despite the challenges.

**Highlights:**

- COVID-19 has impacted cardiac rehabilitation (CR) delivery around the globe.
- In this cross-sectional study, a survey was completed by 1062 (18.3%) CR programs from 70 (63.1%) countries.
- The pandemic has resulted in cessation of ∼75% of CR programs, with others ceasing initiation of new patients, reducing components delivered, and/or changing of mode delivery with little opportunity for planning and training.
- There is also significant psychosocial and economic impact on CR providers.
- Alternative CR model (e.g. home-based, virtual) reimbursement advocacy is needed, to ensure safe, accessible secondary prevention delivery.

## Introduction

The coronavirus disease 2019 (COVID-19) is an infectious disease with high transmissibility caused by severe acute respiratory syndrome coronavirus 2 (SARS-CoV-2), [1] which was declared a pandemic on March 11, 2020 by the World Health Organization (WHO). COVID-19 disproportionately affects older adults, and there is a higher case fatality rate in those with underlying conditions such as cardiovascular diseases (CVD) [2,3]. In addition to respiratory effects, COVID-19 has negative cardiovascular effects as well [4,5]. COVID-19 has placed tremendous pressures on health care organizations around the world.

CVDs are among the most prevalent non-communicable diseases worldwide [6]. Cardiac rehabilitation (CR) is recommended as the standard of care for secondary prevention of CVD [7,8] as it significantly reduces morbidity and mortality [9]. Although models vary, CR is generally delivered in clinical settings [10], involving semi-weekly visits for exercise and education/counselling sessions over several months, for a median of 24 sessions globally [11]. These in-person visits therefore, if continued during COVID-19, carry a high risk of exposure.

Government response and restrictions to the COVID-19 pandemic have varied widely, but include limits to non-essential health care services, including non-urgent outpatient appointments. As a result, the delivery of CR has been impacted. Early information suggests many CR programs globally have closed or switched to virtual delivery [12-21]. This has negative impacts on patients, who are fearful of, or cannot access, cardiac care, adding to this health crisis [22-24]. In addition, CR providers are experiencing occupational health, and economic threats [25]. Therefore, the objective of this study was to investigate impacts of COVID-19 on CR delivery around the globe, including impacts on providers and patients.

## Methods

### Design and Procedure

This research was cross-sectional in design. This work was undertaken by the International Council of Cardiovascular Prevention and Rehabilitation (ICCPR; globalcardiacrehab.com); York University’s Office of Research Ethics (Toronto, Canada) approved secondary use of the data for publication purposes. Participation in the survey constituted voluntary consent to participate. Responses were confidential.

All 40 ICCPR member associations and 10 “friend” members were contacted, and requested to circulate the survey to their members and any other programs of which they were aware in their regions. Contacts of the investigators were also approached to circulate and complete the survey. Potential respondents were also recruited from the attendance list of the ICCPR webinar on COVID-19 and CR [26], which was attended by more than 300 participants.

Data collection occurred from March to June 2020 via online survey administered through REDCap (English). The original survey was not available within China. Therefore, the survey was translated (Simplified Chinese) by co-author XL, a bilingual nurse with CR expertise, and disseminated through Sojump, a professional online survey platform.

### Sample

The sample comprised CR programs around the world, with programs defined as those that offer (1) initial assessment, (2) structured exercise, and (3) at least one other strategy to control risk factors. We asked the CR program manager to complete the survey where possible.

Countries were categorized by WHO region [27]. To ultimately understand representativeness of the sample, the availability of CR by country and programs per country was ascertained from ICCPR’s first global audit of CR (2016/2017) [28]. Through that process, it was estimated that there were 5,813 programs in 111/203 countries with CR internationally (note in the global audit only a random subsample of programs was recruited in the United States, but all programs were emailed for this study) [28]. Information from key informants and respondents were used to update these numbers where available.

### Measures

A 33-item questionnaire was developed by the investigators to examine the objectives of this study (Online Appendix 1). Non-COVID-related questionnaire items were based on ICCPR’s previous CR program survey [18]. The questionnaire was divided into three sections: (1) CR program characteristics and impacts of COVID-19, (2) barriers and facilitators to delivering CR programs virtually, and (3) COVID-related impacts on staff and patients. The items had forced-choice (some were select all that apply) and open-ended response options, and skip-logic was used to get more detail where applicable. Input from CR providers was solicited, with minor changes made to questions and wording prior to launch of the survey.

Respondents were asked to report their country and the date. To contextualize responses, information on COVID-19 cases at the time of survey completion in each country was extracted from the website “Our World in Data” [29] and confirmed with data available on each country’s government website. The Government Response Stringency Index - information on 9 common policy responses that governments have taken to respond to the pandemic such as workplace closures and stay-at-home requirements (rescaled to a value from 0 to 100, with 100 = strictest responses) – was also extracted from this website for each country, with information corresponding to the mean date of survey completion by respondents in each country.

### Data Analysis

Data were exported from REDCap and Sojump to SPSS version 26, where all the analysis was performed. All initiated surveys that had at least one response were included. The number of responses for each question varied due to missing data (e.g., respondent did not answer a question due to inapplicability, skip logic, or decided to not answer for other reasons); for descriptive analyses, percentages were computed using the number of valid responses for the specific item as the denominator.

Descriptive statistics (e.g., frequency with percentage) were applied for all closed-ended items in the survey. All open-ended responses were coded.

## Results

### Respondents

Overall, 1062 responses were received, of which 1044 (98.3%) identified their country. As shown in Table 1, responses were received from 70 (34.5%) of the approximately 203 countries in the world (covering all 6 WHO regions; response rate by region ranged from 14.1-100.0%; median number of responding programs per country=98.0; Q25-75=36.0-108.0), or 63.1% of the 111 countries identified to have any CR programs in ICCPR’s global audit [28]. Given the estimated number of programs globally^28^ and updates gleaned through this process (no additional countries were determined to have CR, but 13 countries had increases in the number of programs since ICCPR’s 2016/17 audit, with a total of 60 additional programs globally; Table 1), responses were received from 18.3% of the estimated 5,813 programs worldwide.

**Table 1:**
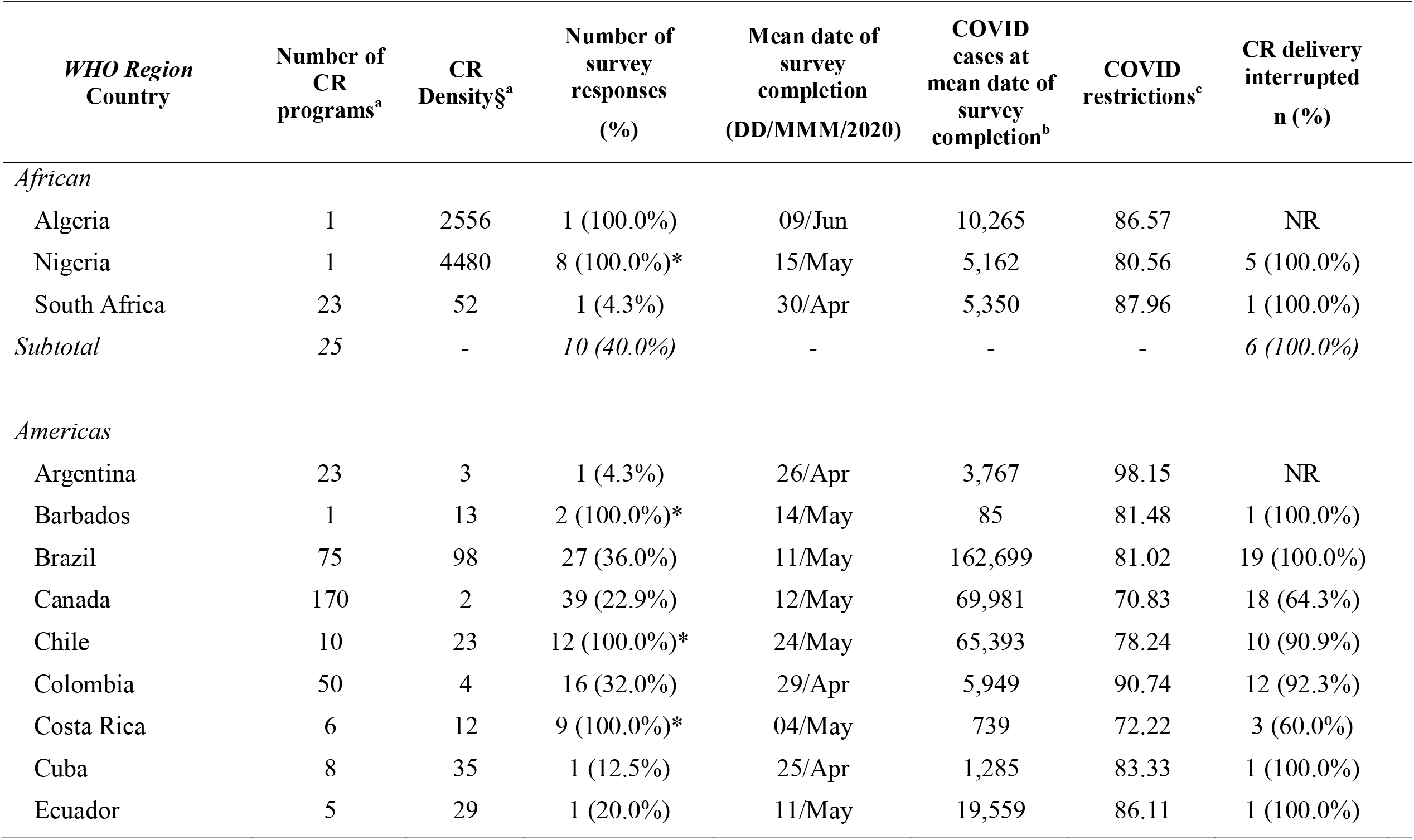
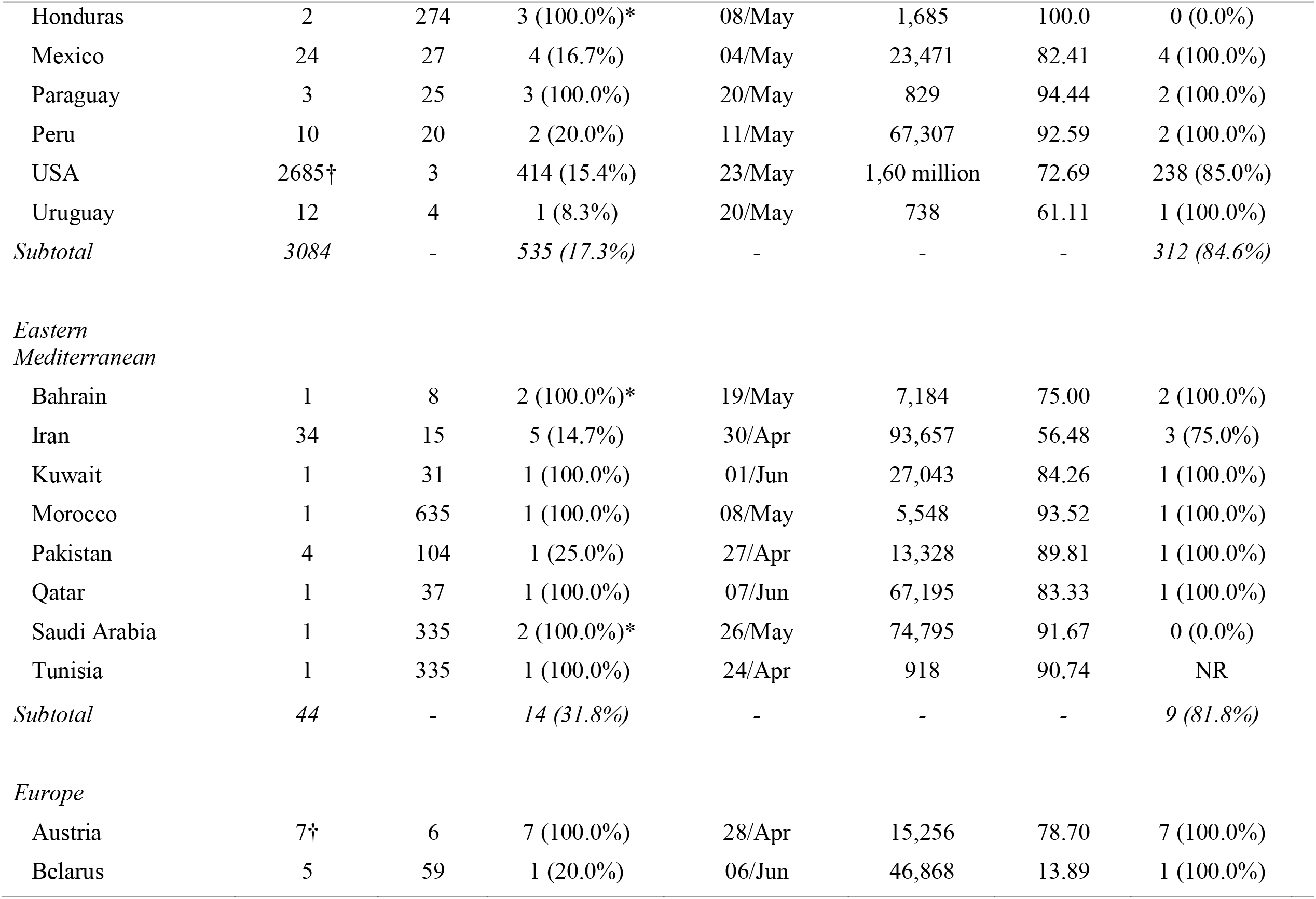
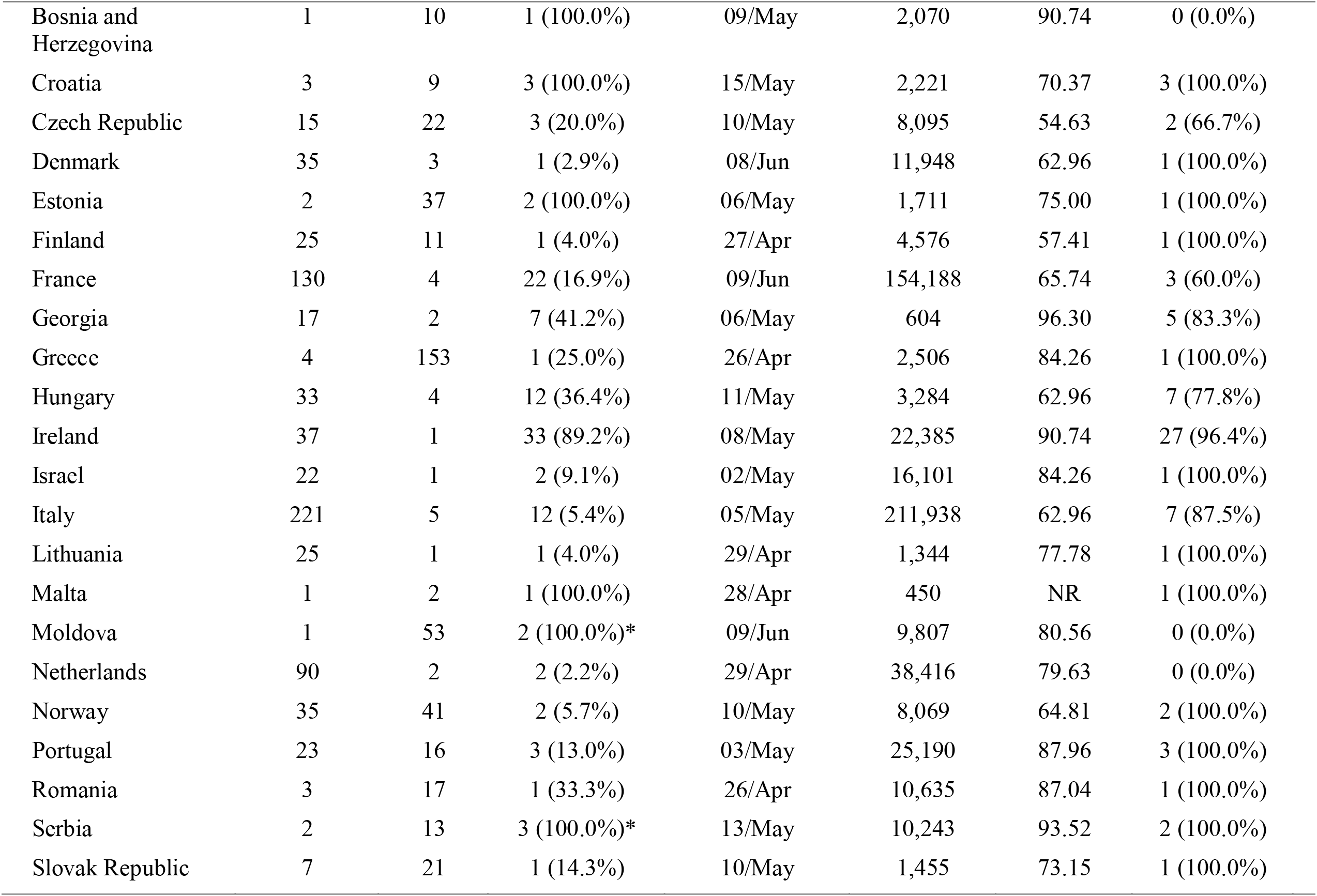
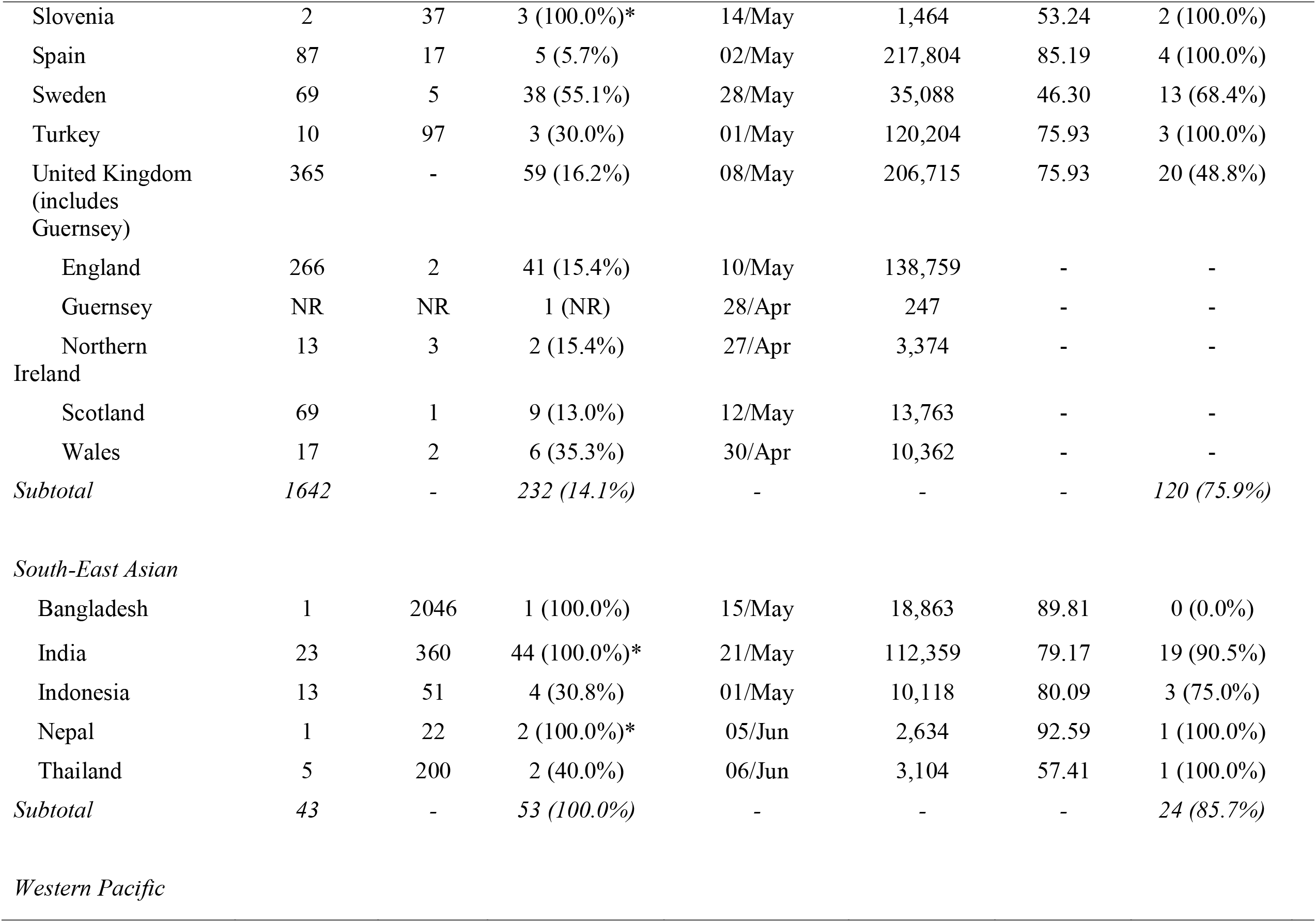
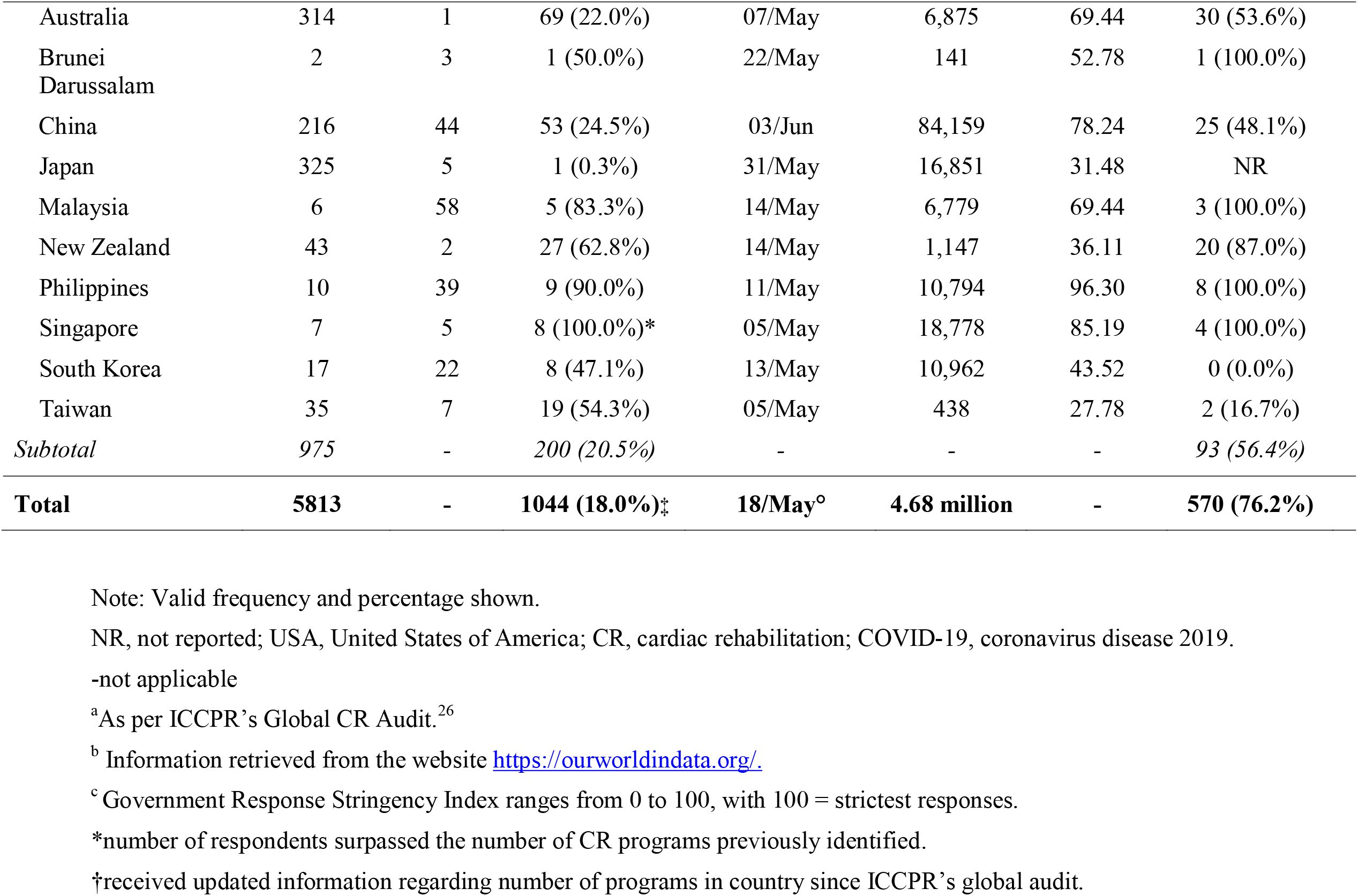
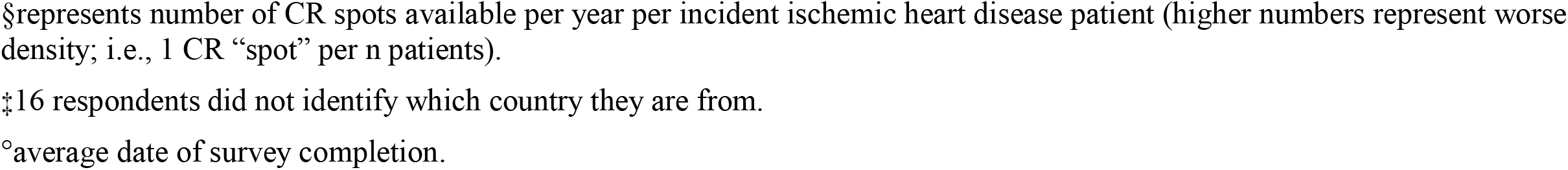
CR availability by country (N=111), survey response and COVID-19 burden

| *WHO Region*<br>Country | Number of<br>CR<br>programs<sup>a</sup> | CR<br>Density§<sup>a</sup> | Number of<br>survey<br>responses<br>(%) | Mean date of<br>survey<br>completion<br>(DD/MMM/2020) | COVID<br>cases at<br>mean date of<br>survey<br>completion<sup>b</sup> | COVID<br>restrictions<sup>c</sup> | CR delivery<br>interrupted<br>n (%) |
| --- | --- | --- | --- | --- | --- | --- | --- |
| *African* |  |  |  |  |  |  |  |
| Algeria | 1 | 2556 | 1 (100.0%) | 09/Jun | 10,265 | 86.57 | NR |
| Nigeria | 1 | 4480 | 8 (100.0%)* | 15/May | 5,162 | 80.56 | 5 (100.0%) |
| South Africa | 23 | 52 | 1 (4.3%) | 30/Apr | 5,350 | 87.96 | 1 (100.0%) |
| *Subtotal* | 25 | - | 10 (40.0%) | - | - | - | 6 (100.0%) |
| *Americas* |  |  |  |  |  |  |  |
| Argentina | 23 | 3 | 1 (4.3%) | 26/Apr | 3,767 | 98.15 | NR |
| Barbados | 1 | 13 | 2 (100.0%)* | 14/May | 85 | 81.48 | 1 (100.0%) |
| Brazil | 75 | 98 | 27 (36.0%) | 11/May | 162,699 | 81.02 | 19 (100.0%) |
| Canada | 170 | 2 | 39 (22.9%) | 12/May | 69,981 | 70.83 | 18 (64.3%) |
| Chile | 10 | 23 | 12 (100.0%)* | 24/May | 65,393 | 78.24 | 10 (90.9%) |
| Colombia | 50 | 4 | 16 (32.0%) | 29/Apr | 5,949 | 90.74 | 12 (92.3%) |
| Costa Rica | 6 | 12 | 9 (100.0%)* | 04/May | 739 | 72.22 | 3 (60.0%) |
| Cuba | 8 | 35 | 1 (12.5%) | 25/Apr | 1,285 | 83.33 | 1 (100.0%) |
| Ecuador | 5 | 29 | 1 (20.0%) | 11/May | 19,559 | 86.11 | 1 (100.0%) |
| Honduras | 2 | 274 | 3 (100.0%)* | 08/May | 1,685 | 100.0 | 0 (0.0%) |
| Mexico | 24 | 27 | 4 (16.7%) | 04/May | 23,471 | 82.41 | 4 (100.0%) |
| Paraguay | 3 | 25 | 3 (100.0%) | 20/May | 829 | 94.44 | 2 (100.0%) |
| Peru | 10 | 20 | 2 (20.0%) | 11/May | 67,307 | 92.59 | 2 (100.0%) |
| USA | 2685† | 3 | 414 (15.4%) | 23/May | 1,60 million | 72.69 | 238 (85.0%) |
| Uruguay | 12 | 4 | 1 (8.3%) | 20/May | 738 | 61.11 | 1 (100.0%) |
| *Subtotal* | *3084* | - | *535 (17.3%)* | - | - | - | *312 (84.6%)* |
| *Eastern Mediterranean* |  |  |  |  |  |  |  |
| Bahrain | 1 | 8 | 2 (100.0%)* | 19/May | 7,184 | 75.00 | 2 (100.0%) |
| Iran | 34 | 15 | 5 (14.7%) | 30/Apr | 93,657 | 56.48 | 3 (75.0%) |
| Kuwait | 1 | 31 | 1 (100.0%) | 01/Jun | 27,043 | 84.26 | 1 (100.0%) |
| Morocco | 1 | 635 | 1 (100.0%) | 08/May | 5,548 | 93.52 | 1 (100.0%) |
| Pakistan | 4 | 104 | 1 (25.0%) | 27/Apr | 13,328 | 89.81 | 1 (100.0%) |
| Qatar | 1 | 37 | 1 (100.0%) | 07/Jun | 67,195 | 83.33 | 1 (100.0%) |
| Saudi Arabia | 1 | 335 | 2 (100.0%)* | 26/May | 74,795 | 91.67 | 0 (0.0%) |
| Tunisia | 1 | 335 | 1 (100.0%) | 24/Apr | 918 | 90.74 | NR |
| *Subtotal* | *44* | - | *14 (31.8%)* | - | - | - | *9 (81.8%)* |
| *Europe* |  |  |  |  |  |  |  |
| Austria | 7† | 6 | 7 (100.0%) | 28/Apr | 15,256 | 78.70 | 7 (100.0%) |
| Belarus | 5 | 59 | 1 (20.0%) | 06/Jun | 46,868 | 13.89 | 1 (100.0%) |
| Bosnia and Herzegovina | 1 | 10 | 1 (100.0%) | 09/May | 2,070 | 90.74 | 0 (0.0%) |
| Croatia | 3 | 9 | 3 (100.0%) | 15/May | 2,221 | 70.37 | 3 (100.0%) |
| Czech Republic | 15 | 22 | 3 (20.0%) | 10/May | 8,095 | 54.63 | 2 (66.7%) |
| Denmark | 35 | 3 | 1 (2.9%) | 08/Jun | 11,948 | 62.96 | 1 (100.0%) |
| Estonia | 2 | 37 | 2 (100.0%) | 06/May | 1,711 | 75.00 | 1 (100.0%) |
| Finland | 25 | 11 | 1 (4.0%) | 27/Apr | 4,576 | 57.41 | 1 (100.0%) |
| France | 130 | 4 | 22 (16.9%) | 09/Jun | 154,188 | 65.74 | 3 (60.0%) |
| Georgia | 17 | 2 | 7 (41.2%) | 06/May | 604 | 96.30 | 5 (83.3%) |
| Greece | 4 | 153 | 1 (25.0%) | 26/Apr | 2,506 | 84.26 | 1 (100.0%) |
| Hungary | 33 | 4 | 12 (36.4%) | 11/May | 3,284 | 62.96 | 7 (77.8%) |
| Ireland | 37 | 1 | 33 (89.2%) | 08/May | 22,385 | 90.74 | 27 (96.4%) |
| Israel | 22 | 1 | 2 (9.1%) | 02/May | 16,101 | 84.26 | 1 (100.0%) |
| Italy | 221 | 5 | 12 (5.4%) | 05/May | 211,938 | 62.96 | 7 (87.5%) |
| Lithuania | 25 | 1 | 1 (4.0%) | 29/Apr | 1,344 | 77.78 | 1 (100.0%) |
| Malta | 1 | 2 | 1 (100.0%) | 28/Apr | 450 | NR | 1 (100.0%) |
| Moldova | 1 | 53 | 2 (100.0%)* | 09/Jun | 9,807 | 80.56 | 0 (0.0%) |
| Netherlands | 90 | 2 | 2 (2.2%) | 29/Apr | 38,416 | 79.63 | 0 (0.0%) |
| Norway | 35 | 41 | 2 (5.7%) | 10/May | 8,069 | 64.81 | 2 (100.0%) |
| Portugal | 23 | 16 | 3 (13.0%) | 03/May | 25,190 | 87.96 | 3 (100.0%) |
| Romania | 3 | 17 | 1 (33.3%) | 26/Apr | 10,635 | 87.04 | 1 (100.0%) |
| Serbia | 2 | 13 | 3 (100.0%)* | 13/May | 10,243 | 93.52 | 2 (100.0%) |
| Slovak Republic | 7 | 21 | 1 (14.3%) | 10/May | 1,455 | 73.15 | 1 (100.0%) |
| Slovenia | 2 | 37 | 3 (100.0%)* | 14/May | 1,464 | 53.24 | 2 (100.0%) |
| Spain | 87 | 17 | 5 (5.7%) | 02/May | 217,804 | 85.19 | 4 (100.0%) |
| Sweden | 69 | 5 | 38 (55.1%) | 28/May | 35,088 | 46.30 | 13 (68.4%) |
| Turkey | 10 | 97 | 3 (30.0%) | 01/May | 120,204 | 75.93 | 3 (100.0%) |
| United Kingdom<br>(includes<br>Guernsey) | 365 | - | 59 (16.2%) | 08/May | 206,715 | 75.93 | 20 (48.8%) |
| England | 266 | 2 | 41 (15.4%) | 10/May | 138,759 | - | - |
| Guernsey | NR | NR | 1 (NR) | 28/Apr | 247 | - | - |
| Northern<br>Ireland | 13 | 3 | 2 (15.4%) | 27/Apr | 3,374 | - | - |
| Scotland | 69 | 1 | 9 (13.0%) | 12/May | 13,763 | - | - |
| Wales | 17 | 2 | 6 (35.3%) | 30/Apr | 10,362 | - | - |
| *Subtotal* | *1642* | *-* | *232 (14.1%)* | *-* | *-* | *-* | *120 (75.9%)* |
| *South-East Asian* |  |  |  |  |  |  |  |
| Bangladesh | 1 | 2046 | 1 (100.0%) | 15/May | 18,863 | 89.81 | 0 (0.0%) |
| India | 23 | 360 | 44 (100.0%)* | 21/May | 112,359 | 79.17 | 19 (90.5%) |
| Indonesia | 13 | 51 | 4 (30.8%) | 01/May | 10,118 | 80.09 | 3 (75.0%) |
| Nepal | 1 | 22 | 2 (100.0%)* | 05/Jun | 2,634 | 92.59 | 1 (100.0%) |
| Thailand | 5 | 200 | 2 (40.0%) | 06/Jun | 3,104 | 57.41 | 1 (100.0%) |
| *Subtotal* | *43* | *-* | *53 (100.0%)* | *-* | *-* | *-* | *24 (85.7%)* |
| *Western Pacific* |  |  |  |  |  |  |  |
| Australia | 314 | 1 | 69 (22.0%) | 07/May | 6,875 | 69.44 | 30 (53.6%) |
| Brunei Darussalam | 2 | 3 | 1 (50.0%) | 22/May | 141 | 52.78 | 1 (100.0%) |
| China | 216 | 44 | 53 (24.5%) | 03/Jun | 84,159 | 78.24 | 25 (48.1%) |
| Japan | 325 | 5 | 1 (0.3%) | 31/May | 16,851 | 31.48 | NR |
| Malaysia | 6 | 58 | 5 (83.3%) | 14/May | 6,779 | 69.44 | 3 (100.0%) |
| New Zealand | 43 | 2 | 27 (62.8%) | 14/May | 1,147 | 36.11 | 20 (87.0%) |
| Philippines | 10 | 39 | 9 (90.0%) | 11/May | 10,794 | 96.30 | 8 (100.0%) |
| Singapore | 7 | 5 | 8 (100.0%)* | 05/May | 18,778 | 85.19 | 4 (100.0%) |
| South Korea | 17 | 22 | 8 (47.1%) | 13/May | 10,962 | 43.52 | 0 (0.0%) |
| Taiwan | 35 | 7 | 19 (54.3%) | 05/May | 438 | 27.78 | 2 (16.7%) |
| *Subtotal* | *975* | *-* | *200 (20.5%)* | *-* | *-* | *-* | *93 (56.4%)* |
| **Total** | **5813** | **-** | **1044 (18.0%)‡** | **18/May°** | **4.68 million** | **-** | **570 (76.2%)** |

Note: Valid frequency and percentage shown.
NR, not reported; USA, United States of America; CR, cardiac rehabilitation; COVID-19, coronavirus disease 2019.
-not applicable
<sup>a</sup>As per ICCPR's Global CR Audit.<sup>26</sup>
<sup>b</sup> Information retrieved from the website https://ourworldindata.org/.
<sup>c</sup> Government Response Stringency Index ranges from 0 to 100, with 100 = strictest responses.
\*number of respondents surpassed the number of CR programs previously identified.
‡received updated information regarding number of programs in country since ICCPR's global audit.
§represents number of CR spots available per year per incident ischemic heart disease patient (higher numbers represent worse density; i.e., 1 CR “spot” per n patients).
‡16 respondents did not identify which country they are from.
°average date of survey completion.

In regards to location, 392 (37.4%) CR programs were located in a community hospital, 364 (34.7%) in a referral center, quaternary or tertiary facility, and/or academic center, 46 (4.4%) in a rehabilitation hospital or residential facility, and 70 (6.7%) in other locations, including private clinics, universities, and fitness centers. With regards to reimbursement, 690 (66.4%) programs have their services covered by the government, 557 (52.8%) programs’ services were paid directly by patients, and 484 (47.5%) were paid by health insurance (many programs had multiple sources).

Table 1 presents number of cases and stringency index by country. The COVID-19 restrictions that had taken place in their area at any time before respondents completed the survey were as follows: 961 (80.2%) reported that gatherings over a certain number of people were prohibited, 947 (79.4%) reported that anyone potentially exposed to COVID-19 was under quarantine or self-isolation, 891 (75.3%) reported schools were closed, 859 (72.1%) reported hospitals were not allowing visitors, 848 (71.7%) reported hospitals were not doing elective surgeries, 840 (71.2%) reported all but essential services/ businesses were closed, 797 (67.0%) reported that people had to wear masks when physical distancing was not possible, 649 (54.4%) reported travel within their region was restricted, and 633 (54.4%) reported the enforcement of physical distancing with fines and other means.

### Impact of COVID-19 on CR Program Delivery

Overall, 106 (14.4%) programs had suspected or positive COVID-19 patients (see Table 1 for COVID-19 cases by region). At the time of survey completion, 367 (49.1%) programs reported they had completely stopped CR delivery for the COVID-19 pandemic, and 203 (27.1%) stopped for a period but had already resumed services (mean 8.3±2.8 [standard deviation] weeks); 178 (23.8%) programs did not stop (see Table 1 for this information by country). As shown, Nigeria has the poorest CR density, and all programs closed. Of those that stopped for any amount of time, 363 (30.0%) made no other arrangements to provide patient care, and the 202 (16.7%) that did describe the following: home-based CR/ telehealth, online consultations (with respondents reporting WeChat, MS Teams, and Zoom), phone or email consultations, and education offered more often via phone, online or via postal mail to patients. During COVID-19, 202 (42.3%) programs were only treating existing patients, while all others were still accepting new patients; programs reported having the capacity to serve a median of 20.0 (Q25-27 =10.0-40.0) patients / month pre-COVID, and 3.0 (Q25-75=0.0-15.0) during. The most common program adaptations were: reducing the number of program elements offered (n=228, 19.5%), deferring graduation until post-program assessments could be completed (n=120; 10.3%), shortening the program duration (n=95, 8.2%), and graduating patients more quickly (n=85, 7.3%). On the other hand, 177 (15.1%) respondents reported adapting all program elements to retain service levels (see Table 2 for adaptations by WHO region).

**Table 2:** Cardiac Rehabilitation Program Adaptations due to COVID-19 by World Health Organization Region

|  | WHO region |  |  |  |  |  | Total |
| --- | --- | --- | --- | --- | --- | --- | --- |
|  | Africa | Americas | Eastern Mediterranean | Europe | South-East Asian | Western Pacific |  |
| Reducing some elements offered | 3 (30.0%) | 110 (20.6%) | 3 (21.4%) | 57 (24.6%) | 10 (18.9%) | 43 (27.0%) | 228 (19.5%) |
| Only treating existing patients | 2 (33.3%) | 100 (41.3%) | 4 (57.1%) | 51 (46.8%) | 14 (63.6%) | 29 (33.3%) | 202 (17.3%) |
| Deferring graduation until post-program assessments can be completed | 2 (20.0%) | 71 (13.3%) | 2 (14.3%) | 21 (9.1%) | 3 (5.7%) | 19 (12.3%) | 120 (10.3%) |
| Shortening the program duration | 3 (30.0%) | 46 (8.6%) | 1 (7.1%) | 18 (7.8%) | 4 (7.5%) | 22 (14.1%) | 95 (8.2%) |
| Graduating patients more quickly | 2 (20.0%) | 57 (10.7%) | 1 (7.1%) | 7 (3.0%) | 2 (3.8%) | 14 (9.2%) | 85 (7.3%) |
| None of the above; We are adapting all elements to retain service levels. | 2 (20.0%) | 91 (17.0%) | 6 (42.9%) | 31 (13.4%) | 12 (22.6%) | 35 (21.6%) | 177 (15.1%) |

Note 1: Valid frequency and percentage shown; respondents instructed to check all that apply.
Note 2: Total does not sum the values from WHO regions because some respondents did not report their country.
WHO, World Health Organization.

Table 3 displays the components that are delivered by CR programs, and continue to be provided during the COVID-19 pandemic. As shown, most impacted was supervised exercise training, resistance training, inclusion of family / informal caregivers, end of program re-assessment, and functional capacity testing. Indeed, of the programs providing services or without COVID-19 restrictions, 119 (14.5%) had changed the type of functional capacity test they use. Of these, 30 (26.3%) were performing functional capacity testing remotely, most commonly: a walk test (n=19; 1.6%), step test (n=6; 0.5%); 15 (1.3%) cited other tests, such as the Duke Activity Status Index and timed sit-to-stand.

**Table 3:** CR components provided in the programs, and that continued to be provided during the COVID-19 pandemic

|  | **Offered and**<br>**continue to be**<br>**offered during**<br>**COVID-19** | **Offered, but**<br>**not during**<br>**COVID-19** | **Never**<br>**Offered** |
| --- | --- | --- | --- |
| Initial assessment | 371 (45.3%) | 390 (47.6%) | 58 (7.1%) |
| Individual consultation with a doctor or nurse | 356 (44.0%) | 335 (41.4%) | 118 (14.6%) |
| Exercise stress test | 144 (18.3%) | 288 (36.5%) | 357 (45.2%) |
| Other functional capacity test | 166 (20.9%) | 438 (55.2%) | 189 (23.8%) |
| Exercise prescription | 375 (46.0%) | 386 (47.3%) | 55 (6.7%) |
| Supervised exercise training | 208 (25.5%) | 521 (63.9%) | 86 (10.6%) |
| Resistance training | 224 (27.9%) | 480 (59.8%) | 99 (12.3%) |
| Patient education | 445 (54.4%) | 331 (40.5%) | 42 (5.1%) |
| Management of cardiovascular risk factors | 436 (54.6%) | 320 (40.1%) | 42 (5.3%) |
| Prescription / titration of secondary prevention medications | 271 (33.9%) | 250 (31.3%) | 278 (34.8%) |
| Nutrition counseling | 352 (43.4%) | 377 (46.5%) | 82 (10.1%) |
| Psychological counseling | 298 (37.3%) | 301 (37.7%) | 199 (24.9%) |
| Smoking cessation sessions/classes | 199 (25.1%) | 344 (43.3%) | 251 (31.6%) |
| Sexual counselling | 134 (17.2%) | 219 (28.2%) | 424 (54.6%) |
| Vocational counseling / support for return-to-work | 166 (21.3%) | 238 (30.6%) | 374 (48.1%) |
| Stress management / Relaxation techniques | 260 (32.6%) | 370 (46.4%) | 168 (21.1%) |
| Alternative forms of exercise (e.g. yoga, dance, or tai chi) | 94 (12.1%) | 174 (22.4%) | 508 (65.5%) |
| Inclusion of family / informal caregivers | 179 (22.5%) | 465 (58.6%) | 150 (18.9%) |
| End of program re-assessment | 245 (30.4%) | 449 (55.7%) | 112 (13.9%) |
| Communication of patient assessment results with their primary care provider | 322 (40.5%) | 342 (43.0%) | 132 (16.6%) |
| Follow-up after outpatient program | 198 (25.3%) | 294 (37.5%) | 292 (37.2%) |
| Maintenance program | 147 (18.7%) | 404 (51.3%) | 236 (30.0%) |
| Other (e.g. telephone sessions, mindfulness sessions) | 36 (10.8%) | 30 (9.0%) | 268 (22.2%) |

Note: Valid frequency and percentage shown.
CR, cardiac rehabilitation.

Of those that continued providing services, the COVID-19 pandemic had impacted the way exercise is prescribed for 306 (37.5%) programs (e.g., virtual consultations with no structured supervised exercise program), and monitored for 226 (27.8%; e.g., focus on rating of perceived exertion rather than target heart rate, all done virtually). Where patients were encouraged to exercise at home or in their community, the following changes were made to manage patient symptoms, reporting or concerns: education about symptoms enhanced, regular virtual (phone and online) consultations with CR staff were added, and patients were encouraged to directly contact their physician if symptoms were experienced.

Furthermore, of programs continuing to provide services, in 111 (13.6%) programs, the type of provider delivering CR had changed, and in 122 (15.0%) patients have had to interact with a different provider (e.g., mainly one professional interacting with patients – mostly nurses, as access to allied health care providers ceased due to COVID-19).

Online Appendix 2 provides details of patient education delivery modes and providers, in light of the COVID-19 pandemic. As shown, there was a drastic reduction in exposure to all disciplines. Modes of delivery were chiefly phone, email, online resources, and mail; live video was only exploited in approximately 20% of programs. For those that offered it, education session frequency before COVID-19 was 8.7±8.4 / month / patient (mean of 50.1±47.5 minutes / session), dropping to 4.1±7.5 (mean of 20.2±26.0 minutes / session) during the pandemic.

### Alternative / Remote Delivery of CR due to COVID-19

Reimbursement and delivery of alternative models (e.g., home-based) is shown in Table 4, including the proportion of patients served (shown before COVID by country elsewhere).^30^ Of those delivering alternative models, 226 (31.7%) programs offered low-tech home-based (e.g., landlines, mail only), 175 (24.5%) offered high-tech home-based, 83 (11.7%) some hybrid format with supervised and unsupervised sessions, 74 (10.4%) offered community-based, and 6 (0.9%) offered “other” models (e.g., livestream).

**Table 4:** Alternative models of CR delivery

|  | **Yes, before**<br>**COVID-19** | **Yes, during**<br>**COVID-19** | **No** |
| --- | --- | --- | --- |
| Reimbursement of alternative models | 102 (12.7%) | 67 (8.3%) | 636 (79.0%) |
| Alternative models offered | 150 (18.5%) | 172 (21.2%) | 488 (60.2%) |
| Proportion of patients served in remote model | 16.5±24.5% | 69.0±37.2% | n/a |
| Perceive program has sufficient capacity to meet need/demand for remote delivery | 142 (49.1%) | 155 (54.0%) | 121 (42.2%) |
| **Forms of Communication** |  |  |  |
| Phone | 178 (62.2%) | 96 (33.6%) | 12 (4.2%) |
| Email | 128 (46.9%) | 88 (32.2%) | 57 (20.9%) |
| Internet webpage | 104 (38.4%) | 99 (36.5%) | 68 (25.1%) |
| Text messages | 93 (34.8%) | 42 (15.7%) | 132 (49.4%) |
| Smartphone app | 52 (20.6%) | 67 (26.6%) | 133 (52.8%) |
| Webcam / videoconference (e.g. education sessions) | 37 (14.2%) | 105 (40.4%) | 118 (45.4%) |
| Other | 14 (15.6%) | 6 (6.7%) | 70 (77.8%) |

Note: frequency and percentage or mean and standard deviation shown.
n/a=not applicable; CR=cardiac rehabilitation

Forms of communication used in remote models are also shown in Table 4. Overall, 182 (62.8%) programs reported barriers to using these communication tools, most commonly: lack of patient access (i.e., patients not having computer with email; n=146, 12.5%), difficulties for the patients (e.g., lack of technical knowledge; n=111, 9.6%), logistical problems such as connectivity issues (n=107, 9.2%), and difficulties for the clinical staff (e.g., being too busy, lack of staff; n=53, 4.6%).

Barriers to delivering CR remotely were: patients did not have the technology to connect with program staff remotely (n=195, 16.8%), lack of equipment /program resources for secure and private remote delivery (n=148, 12.7%), insufficient funding (n=123, 10.5%), insufficient staff (n=118, 10.1%), patients’ risk is too high for unsupervised exercise/safety concerns (n=111, 9.5%), too inefficient (i.e., cannot see as many patients because they do not offer remote program elements in a group, but only individually; n=90, 7.7%), staff need training (n=90, 7.7%), and no policy (n=69, 5.9%); other barriers (n=24, 2.1%) included language proficiency, remote delivery not reimbursed, not enough referrals and hours.

Finally, to increase their capacity to deliver home-based/remote CR services to patients (open-ended item), respondents perceived they would need: time to research and develop the model (including appropriate services for illiterate patients); secure /private means for staff to communicate with patients electronically; equipment to communicate remotely with patients (e.g., multiple laptops with built-in cameras and microphones, headsets); facilities / space (e.g., videoconferencing in private room); home equipment to loan patients including tablets, wearables/activity trackers for exercise monitoring and assessment of all risk factors (e.g., sphygmomanometers/cuffs); cheap and reliable wireless technology to monitor for adverse events remotely (e.g., heart rate and rhythm); reliable and low-cost high-speed internet access for staff and patients (including those living in rural and remote areas); a dedicated multidisciplinary team (including more nurses and exercise physiologists); staff training (by those with experience in remote models), physician champions; administrative staff to facilitate scheduling of virtual sessions; a structured, evidence-based home-based CR program software platform or smartphone app (including that it has capacity to reach groups of patients, that family / informal caregivers can also access it, that it is comprehensive [e.g., includes structured exercise livestreaming, education], and includes tracking diaries with two-way communication; or funding for technology upgrades), with associated assessment tools (including functional capacity) and policies to implement; technology support staff (with time) to train patients (potentially in-person when they are in acute care) and staff to use the remote technology (e.g., zoom) and equipment, and also for database management support; ability to have at least one safe in-person session with each patient to ensure safety and education; financial resources as well as reimbursement of remote model; and patient as well as provider awareness of availability of the remote model (including referrals).

### Impact on CR staff

CR team communication was also impacted. Overall, 295 (25.0%) reported having only ad-hoc meetings as needed, 274 (23.0%) reported they were having routine staff meetings via remote means (e.g., videoconferencing), 137 (11.8%) reported not being able to communicate as much as needed (e.g., staff re-deployed, staff lack home technology), and 88 (7.6%) reported they could not really communicate except via email.

Occupational impacts on staff are displayed in Table 5 by WHO region. Moreover, 114 (15.3%) staff reported feeling stigmatized outside of work for being a healthcare worker (i.e., greater risk of COVID-19 exposure), 275 (36.9%) felt valued for providing care, and 325 (43.6%) felt no stigma (others responded ‘not applicable’). Negative psychosocial impacts that do or could impact their work are shown in Figure 1 (respondents were directed to check all that apply; n=98, 8.4% responded ‘not applicable’ as they have experienced no COVID-19 impacts; n=267, 22.8% of respondents reported no personal suffering due to COVID-19; and n=233, 20.0% have been using the opportunity to catch up on things and learn).

**Table 5:** COVID-19 Impacts by World Health Organization Region

|  | WHO region |  |  |  |  |  | Total |
| --- | --- | --- | --- | --- | --- | --- | --- |
|  | Africa | Americas | Eastern Mediterranean | Europe | South-East Asia | Western Pacific |  |
| Program has had suspected or positive COVID-19 cases | 3 (50.0%) | 47 (12.9%) | 5 (55.6%) | 32 (20.6%) | 3 (10.7%) | 13 (8.0%) | 106 (14.4%) |
| Restrictions on staff work, travel or home life | 4 (66.7%) | 251 (68.0%) | 9 (81.8%) | 98 (62.0%) | 17 (58.6%) | 94 (57.3%) | 481 (64.3%) |
| Concern about delivering CR due to lack of PPE | 5 (83.3%) | 71 (19.5%) | 2 (18.2%) | 45 (28.8%) | 15 (51.7%) | 44 (27.2%) | 182 (24.6%) |
| Felt need to work despite perceived risk for fear of losing job or pay | 4 (66.7%) | 149 (41.0%) | 2 (18.2%) | 43 (27.9%) | 12 (41.4%) | 63 (38.2%) | 276 (37.3%) |
| No paid sick leave | 1 (16.7%) | 74 (20.7%) | 3 (27.3%) | 15 (9.7%) | 12 (41.4%) | 45 (27.3%) | 150 (20.4%) |
| Institution giving staff information so can perform duties safely | 6 (100.0%) | 335 (93.1%) | 10 (100.0%) | 143 (92.9%) | 29 (100.0%) | 163 (98.8%) | 696 (94.6%) |
| Staff Changes |  |  |  |  |  |  |  |
| Re-deployed | 4 (40.0%) | 203 (37.9%) | 7 (50.0%) | 87 (37.5%) | 5 (9.4%) | 41 (25.6%) | 353 (30.2%) |
| Reduced hours | 4 (40.0%) | 163 (30.5%) | 4 (28.6%) | 16 (6.9%) | 4 (7.5%) | 21 (13.5%) | 215 (18.5%) |
| Some staff have had to practice somewhat outside their scope | 3 (30.0%) | 80 (15.0%) | 7 (50.0%) | 41 (17.7%) | 4 (7.5%) | 21 (13.5%) | 159 (13.7%) |
| Laid off temporarily | 1 (10.0%) | 96 (17.9%) | 4 (28.6%) | 21 (9.1%) | 4 (7.5%) | 12 (7.8%) | 138 (11.9%) |
| Pay reductions | 3 (30.0%) | 36 (6.7%) | 4 (28.6%) | 10 (4.3%) | 5 (9.4%) | 13 (8.2%) | 71 (6.1%) |
| Permanently let go | 1 (10.0%) | 14 (2.6%) | 0 (0.0%) | 6 (2.6%) | 1 (1.9%) | 2 (1.0%) | 24 (2.1%) |
| Other | 0 (0.0%) | 24 (4.5%) | 0 (0.0%) | 8 (3.4%) | 2 (3.8%) | 9 (6.0%) | 43 (3.7%) |

Note 1: Frequency and valid percentage shown.
Note 2: Total does not sum the values from WHO regions because some respondents have not reported their countries.
WHO, World Health Organization; PPE, personal protective equipment.

**Figure 1:**
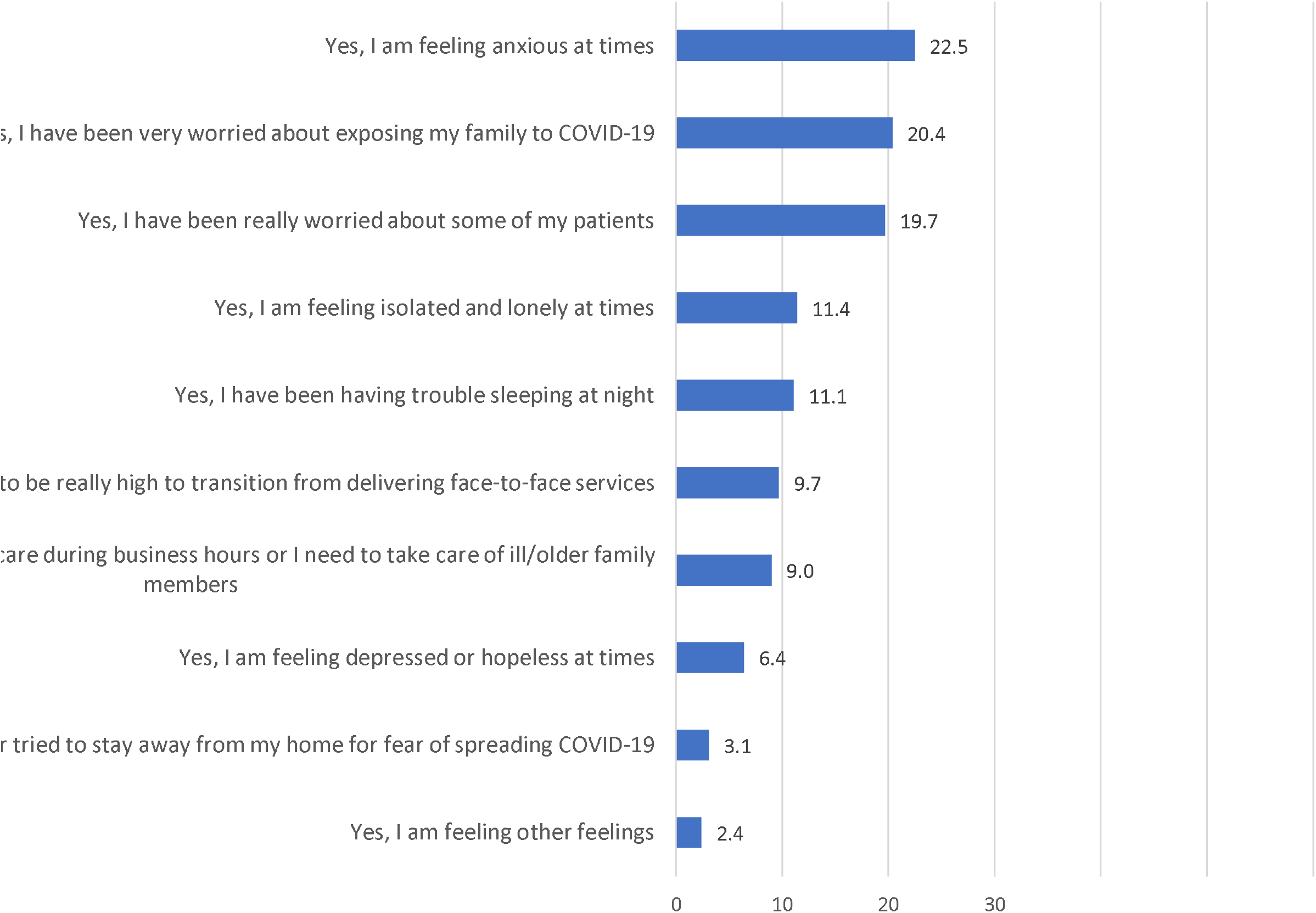
Psychological impacts on CR staff due to COVID-19 Note: Other feelings included anger, fear, frustration, exhaustion, sadness, stress.

When asked whether they had the equipment (e.g., video camera, remote access to hospital servers) and space / privacy to work from home efficiently during business hours, 198 (26.6%) responded affirmatively, 141 (19.0%) responded only for work that does not involve patient care, and 272 (36.6%) reported no (for reasons provided including: no access to work materials and hospital server remotely).

Table 5 also displays whether programs have treated suspected or COVID-19 positive patients by WHO region, and associated concerns. Figure 2 displays perceived risk in contacting COVID-19 through their CR work by country (global mean=30.0%±27.4). Generalized linear mixed models, accounting for country as a higher-order variable, revealed degree of perceived risk was associated with country stringency index (p=0.01), but not number of cases (p=0.80).

**Figure 2:**
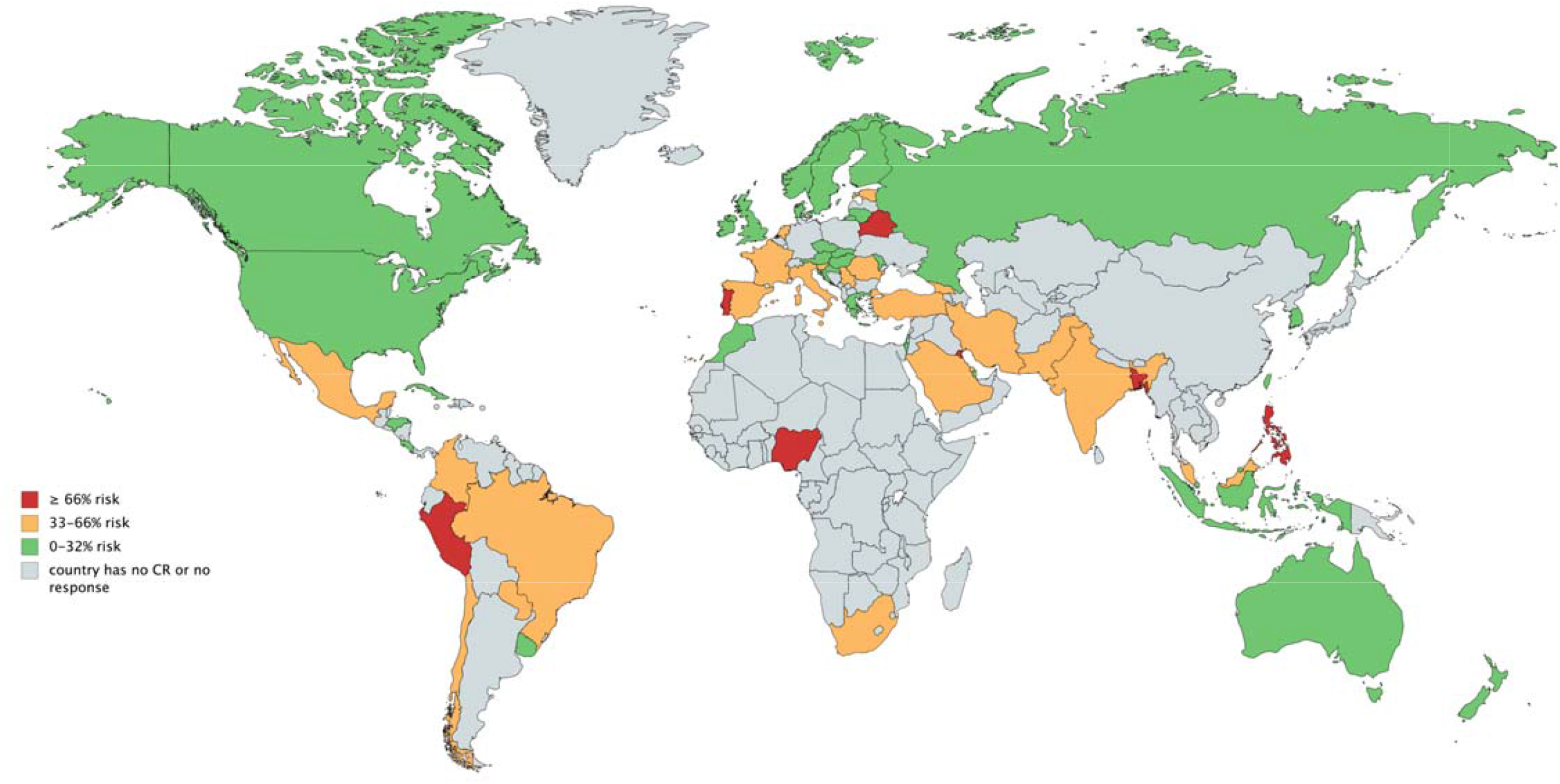
Perceived risk in contacting COVID-19 through their CR work by country

### Impact on Patients

CR staff reported patients contacting them and expressing the following reservations and concerns regarding COVID-19: whether they need to change their exercise prescriptions (n=337, 28.7%), adherence to a heart-healthy diet while concerned about going to grocery stores (n=333, 28.4%), about their mental well-being (n=331; 28.2%), using technology to interact with the program (n=329, 27.9%), questions about medications (n=318, 26.9%), stopping exercise due to a lack of space to exercise in their homes or inability to exercise outside without potential COVID-19 exposure (n=303, 25.7%), questions about how to safely receive CR care without COVID-19 exposure (n=255, 21.4%), their risk of death from COVID-19 due to pre-existing CVD (n=249, 21.2%), and many patients contacted programs about reservations regarding receiving CR care and staff had to let them know they closed down the program temporarily (n=230, 19.6%). On the other hand, 142 (12.2%) reported patients had not really been contacting the program more often due to COVID-19, and 62 (5.3%) reported patients seemed to be avoiding contacting their programs at all, under the assumption that services were suspended.

### Resumption of CR services

Overall, 273 (36.8%) reported that their institution had a policy regarding the circumstances under which regular services could resume (for n=167, 22.5% this was not applicable). Figure 3 illustrates the factors that respondents perceive should form the basis for resumption of face-to-face CR services given COVID-19. Other responses included: number of deaths reduced for 2 consecutive weeks, as well as when government allows gyms to open for business and instructs hospitals to open outpatient services.

**Figure 3:**
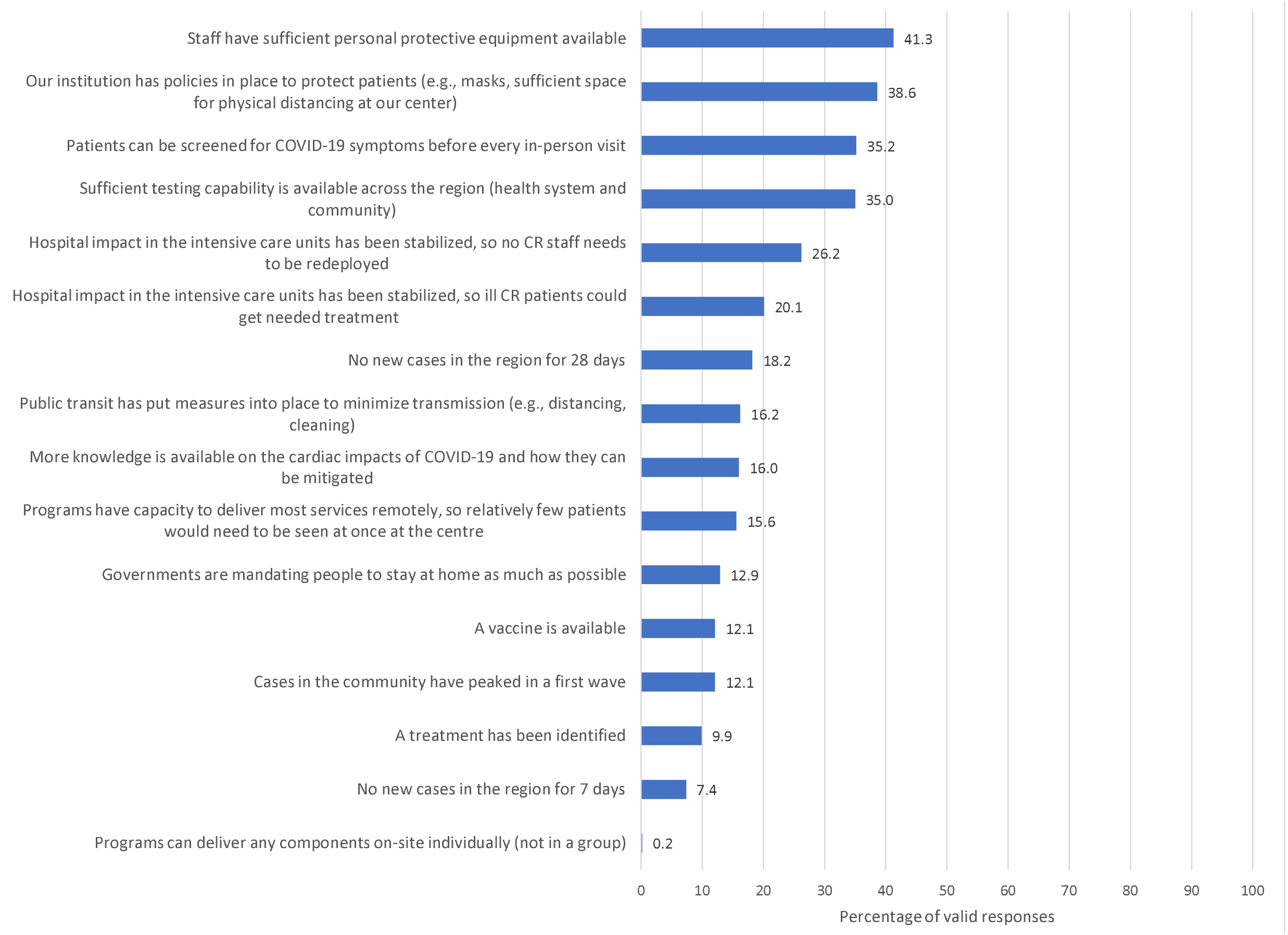
Factors that respondents perceive should form the basis for resumption of face-to-face CR services given COVID-19 Note: respondents instructed to check all that apply.

## Discussion

This first-ever global survey on the impacts of COVID-19 on outpatient rehabilitation has confirmed that the impact of this infectious disease goes well beyond those suffering from it, affecting availability, structure, delivery format, and components of chronic disease care, as well as the mental health of CR providers and patients alike. Given that three-quarters of responding programs reported complete or temporary cessation of services, we estimate COVID-19 has been responsible for the closure of approximately 4400 CR programs worldwide; it is unknown when or if these programs will resume care delivery. Capacity was incredibly low in the programs that remained open (i.e., 3 patients/month, with no new patients) and key risk-reducing components were no longer delivered in more than 60% of on-going programs, including supervised exercise training. This substantial service loss will likely result in greater CVD morbidity and mortality in the coming months and years, further straining already-taxed healthcare systems.

While we must be guided by evidence on transmission and efficacy of protective measures, and monitor progress with regard to vaccination and treatment, results of the study can inform safe resumption of CR services. American [31], British [17] and Canadian [32] CR Associations have published recommendations on resuming face-to-face CR services, which include external and internal (patient, staff, and program) considerations; most of these were also identified as paramount in our study. Specifically, program respondents perceived the bases for resumption of in-person CR services in the context of COVID-19 should chiefly be availability of personal protective equipment (PPE) for staff and patients, screening capacity for COVID-19 symptoms before every in-person visit, sufficient testing capacity, and no CR staff redeployment.

The optimal CR setup once services resume is also unknown. Although considered a good option to continue CR during to the COVID-19 pandemic [15-17], over 60% of programs did not offer any remote model of CR delivery. Many did however, despite the fact that it is not reimbursed in most jurisdictions. Studies have shown that home-based models of CR have similar effects in improving clinical outcomes and quality of life in cardiac patients compared to center-based models [16,33]. Programs that already had remote services significantly increased the proportion of patients served through this modality. Programs are actively trying to continue to deliver CR during this challenging time, but face many barriers, including development of the remote model, as well as the associated policies, and acquiring and learning the new technologies to support this.

To balance both infection risk and risk of an adverse event, some in-person contact at the beginning of the CR program was advocated, following screening and with PPE (indeed one-quarter of respondents were concerned about delivery due to lack of PPE), perhaps at reduced volume to enable physical distancing. Programs have transitioned to using different functional capacity testing methods to inform exercise prescription and different means to monitor exercise. There was great interest in using technology to remotely monitor risk factors, with the goal of loaning required equipment to patients during their program. Most CR components could then be safely delivered through remote means, with the use chiefly of webcam/videoconferencing where resources and supports exist. The significant drop in amount of patient education was concerning; comprehensive, evidence-based virtual patient education is available open access in 8 languages for programs to use [34]. Of note, ICCPR is supporting programs worldwide in transitioning to such lower-risk forms of delivery, by connecting members to share best practices, collating patient and provider resources on our website [35], as well as offering - and promoting other societies’ - free CR webinars to increase provider knowledge. Research is now needed on implementation, patient engagement and outcomes with COVID-adapted CR, as well as cost.

The social restriction, anxiety and occupational impacts resulting from coronaviruses adversely affects the psychological well-being of healthcare workers, and this impact can be substantial and long-lasting [36-38]. Although about a fifth of respondents in this study reported they have been coping “alright” with COVID-19, many are experiencing anxiety, fear of exposing family, loneliness, difficulty sleeping, and stress due to higher workloads (including answering many questions from patients about their concerns). Over a third felt the need to work despite perceived risk, due to fear of losing their job or pay (and one in five had no sick pay). Approximately a third of CR program respondents had been redeployed, rendering it impossible to deliver CR services without staff. Almost 10% have needed to work double-duty, serving as caregivers during business hours. The inequities uncovered by this outbreak [39] are also evidenced in the data from this study, with variation in occupational / economic and psychosocial impacts by WHO region (although inferential tests were not performed due to some small cell sizes).

Caution is warranted in interpreting the findings of this study. First, this was a convenience sample; results may be biased, representing mainly programs that still had some staff working and available to respond to the survey. However, there was good representation from programs in community and academic hospitals alike. We did assume that each response was received from a unique program, however it is possible different staff members from the same program completed a survey, given most respondents were working remotely. Response rate was lower in the Americas and Europe, so generalizability to those regions is more questionable; country representation was high, however. Second, the reliability and validity of the survey is unknown; it was pilot-tested, but responses in a random subsample of programs were not verified against actual delivery. Programs may have responded in a socially-desirable manner, although the survey was confidential. Third, because of small cell sizes in some case, inferential comparisons by WHO region could not be performed. Finally, since the survey was mainly completed in English, some concepts could have been misunderstood by respondents for whom English was not their first language.

## Conclusion

The COVID-19 pandemic has impacted CR programs worldwide, including cessation of services or of initiation of new patients, a decrease in CR components delivered, change of mode delivery without much opportunity for planning and training, as well as psychosocial and economic impact on healthcare providers. Technology is seen as a safe means to ensure cardiac patients receive the care they need during this difficult time, within the context of screening, testing and sufficient PPE. Remote delivery is not often reimbursed, and therefore advocacy is needed if we are to fulfill our mission in supporting patients in secondary prevention, which will ultimately reduce the burden on a strained healthcare system.

## Data Availability

Data is available upon request.

## Acknowledgements

We are grateful to Drs. Karam Turk-Adawi (helped with recruitment for this study) and Marta Supervia for their leadership and collaboration on the first global cardiac rehabilitation audit (https://globalcardiacrehab.com/Global-CR-Program-Survey), which enabled us to undertake this survey expeditiously.

## Author’s Contributions

GLMG and SLG made substantial contributions to the conception and design of the work; the acquisition, analysis, and interpretation of data for the work, and drafting the work. ZX, XL, AM, RG, ASB, SM, CY, JB, PO, and AC supported acquisition of data. All authors: (1) contributed to revising the work critically for important intellectual content and provided final approval of the version to be published; (2) agree to be accountable for all aspects of the work in ensuring that questions related to the accuracy or integrity of any part of the work are appropriately investigated and resolved; and, (3) agreed to be named on the author list, and approved of the full author list.

### Appendix

Appendix 1: The 33-item questionnaire developed by the investigators to examine the objectives of this study

Appendix 2: Patient education mode and providers delivering, under usual practice and during COVID-19

