## Appendix 1 for "Impacts of the COVID-19 Pandemic on Cardiac Rehabilitation Delivery around the World"

**GLOBAL SURVEY OF CARDIOVASCULAR REHABILITATION PROGRAMS: COVID-19 IMPACT**

Instructions: Please answer the series of questions by: (1) checking (🗸) the appropriate box (sometimes one box and other times you will be asked to check as many boxes as apply), (2) typing in an answer, or (3) entering a number, as indicated. The survey items for which you enter numbers are constrained to one value (i.e., you cannot enter a range. If you would like to enter a range, instead enter the midpoint) and will not accept text. You can report a number to up to 1 decimal place if desired. Enter zero (0) only if the answer is none.

Note: CR refers to cardiac rehabilitation.

When answering the questions, please think about the time when the most restrictions were in place due to COVI-19 (if restrictions have eased at the time you are completing this survey)

Be sure to click the “Submit” button when you reach the end of the survey.

**SECTION A: GENERAL INFORMATION**

1. What is today’s date? [date box/calendar]
2. In what country is your cardiac rehabilitation program?

[dropdown menu will be created with all countries identified to have CR in Global CR Survey]

3. Is your cardiac rehabilitation program located within a hospital?
   - Yes – it is in a referral centre/ quarternary / tertiary facility and / or academic centre
   - Yes – it is in a community hospital
   - Yes - it is in a rehabilitation hospital/ residential facility
   - Yes – other
   - No

If no, please specify where your cardiac rehabilitation is located _____________________

4. Who pays for cardiac rehabilitation? (Check all that apply)

- Government / public hospital
- *Patient*
- Private health insurance / private hospital
- *Other (e.g., donations, Foundation, grants)*

Please specify: _____________________

5. What COVID-19 restrictions have been in place at any time in your region? (check all that apply)
   - All but essential services/ businesses are closed
   - Travel within the region is restricted
   - Hospitals are not doing elective surgeries
   - Hospitals do not allow visitors
   - Schools are closed
   - Physical distancing is being enforced with fines or other means
   - Anyone potentially exposed to COVID-19 is under quarantine or self-isolation
   - Gatherings over a certain number of people are prohibited
   - People wear masks when physical distancing is not possible
   - Other (please specify: ____________________________)
   - None of the above

**SECTION B: DETAILS ABOUT YOUR CARDIAC REHABILITATION PROGRAM COMPONENTS**

6. Which of the following elements of cardiac rehabilitation are provided in your program, and continue to be provided during the COVID-19 pandemic? (check one box per row)

|  | **Yes, we offer it and continue to offer during COVID-19** | **Yes, we offer it but not during COVID-19** | **No** |
| --- | --- | --- | --- |
| Initial assessment |  |  |  |
| Individual consultation with a doctor or nurse |  |  |  |
| Exercise stress test |  |  |  |
| Other functional capacity test |  |  |  |
| Exercise prescription |  |  |  |
| Supervised exercise training |  |  |  |
| Resistance training |  |  |  |
| Patient education* |  |  |  |
| Management of cardiovascular risk factors |  |  |  |
| Prescription and/or titration of secondary prevention medications |  |  |  |
| Nutrition counseling |  |  |  |
| Psychological counseling |  |  |  |
| Smoking cessation sessions/classes |  |  |  |
| Sexual counselling |  |  |  |
| Vocational counseling / support for return-to-work |  |  |  |
| Stress management / Relaxation techniques |  |  |  |
| Alternative forms of exercise, such as yoga, dance, or tai chi |  |  |  |
| Inclusion of family / informal caregivers |  |  |  |
| End of program re-assessment |  |  |  |
| Communication of patient assessment results with their primary care provider |  |  |  |
| Follow-up after outpatient program |  |  |  |
| Maintenance program |  |  |  |
| Other** |  |  |  |

*If checked YES for patient education, please go to section “Mode of patient education delivery before and after the COVID-19”, questions 13, 13b, 13c and 13d.

**If checked YES for other, please specify.
Please specify what other components of cardiac rehabilitation are provided in your program: _______________

7. Have you changed the type of functional capacity test used during COVID-19?

- Yes
- No
- Not applicable as we are not providing service during COVID-19 or have had no COVID-19 restrictions

**If checked Yes for question 7**

7a. Are you doing functional capacity testing remotely?

- No
- Yes
- Not applicable as we are not providing service during COVID-19 or have had no COVID-19 restrictions

7b. If yes, what test?

- Step test
- walk test
- other (please specify: ________________________________)

7c. If yes, describe how: ______________________________________________

8. Have you made any changes to how you **prescribe** exercise due to COVID-19, including mode?

- No
- Yes (if yes, please specify: ________________________________________________________)
- Not applicable as we are not providing service during COVID-19 or have had no COVID-19 restrictions

9. Have you made any changes to how you **monitor** exercise due to COVID-19, considering intensity?

- No
- Yes (if yes, please specify: ________________________________________________________)
- Not applicable as we are not providing service during COVID-19 or have had no COVID-19 restrictions

10. If patients are being encouraged to exercise at home and in their community, have you had to make any changes to how you handle patient symptoms, reporting or concerns? If yes, how?

[describe changes or write not applicable]

11. Has the type of provider delivering CR changed due to COVID-19?

- Yes (please specify: ____________________________________________)
- No
- Not applicable as we are not providing service during COVID-19 or have had no COVID-19 restrictions

12. Have patients had to change the CR providers they interact with due to COVID-19 (i.e., interruption to care continuity?)

- Yes (please describe the changes: __________________________________________)
- No
- Not applicable as we are not providing service during COVID-19 or have had no COVID-19 restrictions

**If answered Yes to Patient Education on Question 6,**

**Mode of Patient Education delivered before and after the COVID-19**

a. Which of the following characteristics of patient education intervention are offered in your program and continue to be offered during the COVID-19 pandemic? (check one box per row)

|  | **Yes, we offer it and continue to offer during COVID-19** | **Yes, we offer it but not during COVID-19** | **No** |
| --- | --- | --- | --- |
| One-on-one sessions |  |  |  |
| Group sessions |  |  |  |
| Telephone-based sessions (individual or group) |  |  |  |
| Virtual real-time/live sessions (group) |  |  |  |
| Virtual real-time/live interactive session (one-to-one) |  |  |  |
| Online resources for patients to view/download (asynchronous) |  |  |  |
| E-mailing resources as needed |  |  |  |
| Mailing (postal service) resources as needed |  |  |  |
| Other* |  |  |  |

*If checked OTHER, please specify: __________________.

If education sessions are provided, what is the frequency of sessions each patient receives (i.e. number of sessions):

b_____ times per month before COVID-19

c_____ times per month during COVID-19

If education sessions are provided, what is the duration of each session (in minutes):

d______minutes per session before COVID-19

e_____ minutes per education session during COVID-19

f. If education sessions are provided, who delivers this education (check all that apply, at least one box per row):

|  | **Under usual practice** | **During COVID-19** | **Never delivered education in our program** |
| --- | --- | --- | --- |
| Nurses |  |  |  |
| Exercise leader |  |  |  |
| Medical doctor |  |  |  |
| Healthcare providers from multiple disciplines |  |  |  |
| Other* |  |  |  |

*If checked OTHER, please specify: __________________.

13. Have patients been expressing reservations/concerns related to COVID-19 to staff regularly? (check all that apply)

- Yes, we are getting contacted with questions about how to safely receive CR care, without potential COVID-19 exposure
- Yes, we are getting contacted by patients about how to use technology to interact with the program
- Yes, we are getting contacted by patients with questions about medications
- Yes, we are getting contacted by patients about whether they need to change their exercise prescriptions
- Yes, we are getting contacted by patients who are having to stop their exercise because they have no place to exercise in their homes and they cannot exercise outside without potentially exposing themselves to others
- Yes, we are getting contacted by patients regarding concerns about their mental well-being
- Yes, we are getting contacted by patients about concerns about their risk of death from COVID-19 due to pre-existing heart disease
- Yes, patients are asking us about how to follow a heart healthy diet when they are concerned about going to the grocery store
- Yes, patients were in touch about reservations until we had to let them know we closed down our program temporarily
- Yes, patients are contacting us for other reasons (please specify: ____________________________________)
- No, patients have not really been contacting the program more often due to COVID-19
- No, patients seem to be avoiding contacting our program at all as they assume services are suspended and we will contact them when they resume
- Not applicable

**SECTION C: ALTERNATIVE MODELS OF CR DELIVERY / REMOTE DELIVERY DURING COVID-19**

14. Are alternative cardiac rehabilitation models, such as home-based, reimbursable by government or insurance companies in your region?
    - Yes, always have been
    - Yes, because of COVID-19
    - No
15. Does your cardiac rehabilitation program offer alternative models of program delivery than an on-site program?

- Yes, always have
- Yes, since COVID-19
- No

**If marked Yes to Question 15**

15a. **If marked: yes**, please specify (check all that apply):

- Home-based (low-tech – i.e, landlines, paper)
- Home-based (includes web or Smartphone-based)
- Community-based
- *Hybrid of supervised with home or community-based*
- *Other*

 Please, specify what other alternative model is offered________________

**If Q15a was marked: home–based program, please answer the following questions**:

What percentage of your patients are served in a home-based/remote program? (Enter ‘unknown’ if you do not know) (Please enter a numeric value for the two scenarios)

a.____________ % during usual practice

b. ____________ % during COVID-19

c. Do you perceive your program has sufficient capacity to meet need/demand in the home-based/remote model before COVID-19?

- Yes
- No

d. Is home-based/remote capacity sufficient during COVID-19?

- Yes
- No
- Not applicable as we are not providing service during COVID-19 or have had no COVID-19 restrictions

e. What are the barriers at your program to delivering CR programming to patients remotely? (check all that apply):

- Not enough funding
- Not enough staff
- Too inefficient (i.e., can’t see as many patients because we do not offer remote program elements in a group, but only individually)
- Patients’ risk too high for unsupervised exercise / safety concerns
- Lack of equipment / program resources for secure and private remote delivery
- Patients do not have the tech to connect with program staff remotely
- Staff need training
- No policy
- Other

Please specify the other reason _____________________________________

f. What forms of communication are used with patients in your home-based/remote program or during COVID-19? (check one box per row)

|  | **Yes, we always offer it and continue to offer during COVID-19** | **Yes, we started to offer during COVID-19** | **No** |
| --- | --- | --- | --- |
| Internet webpage (e.g. online resources for patients to view and download) |  |  |  |
| Email (e.g. for forms and resources) |  |  |  |
| Webcam / videoconference (e.g. education sessions) |  |  |  |
| Smartphone app |  |  |  |
| Text messages |  |  |  |
| Telephone (landline) for consultations |  |  |  |
| Other* |  |  |  |

*If checked OTHER, please specify what other form of communication are used:_______________________

g. Does the CR program experience any barriers to using these communication tools?

- *Yes*
- No

If yes: Check all the barriers that apply:

- Logistical problems: i.e., connection
- Lack of patient access (i.e., patients do not have computer with email)
- Difficulty for the clinical staff
  Please specify the difficulties for the clinical staff: ______________
- Difficulty for the patients

Please specify the difficulties for the patients:____________

- Other

Please specify other perceived barriers to communicating with patients via technology: _________________________________________________

h. What do you think you would need to be ready and able to significantly increase your program’s capacity to provide home-based/remote cardiac rehabilitation services to patients?

_________________________________________________________________________________________________________________________________________________________________________________________________________________________________

**SECTION D: DETAILS ABOUT THE IMPACT OF COVID-19 IMPACT**

**ON YOU AND YOUR CR PROGRAM**

16. Has your employer placed any restrictions on your work, travel or home life due to COVID-19?
    - Yes (please specify: ____________________)
    - No
17. Have you felt stigmatized outside of work because you are a healthcare worker who has greater risk of being exposed to COVID-19?
    - Yes
    - No
    - Actually, I feel valued for providing care during this time
    - Not applicable
18. During the COVID-19 pandemic, have you had to stop delivering cardiac rehabilitation programming or was there a period where you were not offering any programming?

- Yes, I had to stop delivering CR
- Yes, I stopped for a period of time. (How long? Please specify in weeks___________________)
- No

**If Yes complete the following questions: 18a – 18e**

18a. If **Yes**, are other arrangements in place to support CR services for your patients during the COVID pandemic?

- Yes (please describe ____________________)
- No

18b How many weeks so far has CR delivery been impacted? ____ weeks

18c Have any staff changes been made? (check all that apply)

- Some staff have been redeployed to COVID-related duties
- Some staff have been laid off temporarily
- Some staff have been permanently let go
- Some staff have had their pay reduced
- Some staff have had their hours reduced
- Some staff have had to practice somewhat outside their scope
- Other (please specify: ___________________________)
- Not applicable

18d During the time of COVID adaptations, is your program:

- Accepting new patients
- Only treating existing patients

18e During the time of COVID adaptations, is your program (check all that apply):

- Shortening the program duration
- Reducing some elements offered
- Graduating patients more quickly
- Deferring graduation until post-program assessments can be completed
- None of the above. We are adapting all elements to retain service levels.

19. How many new **patients** do you have capacity to serve **each month**, in terms of staff and space, if they all completed your program? (Please enter a numeric value; please report the number of new patients that could start your program each month, not the number of patients you treat each year or the number of sessions you offer)

_____________ new **patients per month**

20. How many new cardiac rehabilitation **patients** do you provide service to **each month** during the COVID-19 pandemic? (Please enter a numeric value)

_________ **new patients per month during the COVID-19 pandemic** or

- Not applicable as we are not providing service during COVID-19 or have had no COVID-19 restrictions

21. During the time of COVID-19 adaptations, how is your staff communicating with one another (or are they not)? (check all that apply)
    - We are having routine staff meetings through videoconferencing (e.g., zoom) or another remote medium
    - We are having ad-hoc staff meetings as needed
    - We are not able to communicate as much as needed (e.g., program closed, staff re-deployed, staff lack home technology)
    - We cannot really communicate together as a team, except for email
    - Other (please specify: _____________________________)
    - Not applicable as we are not providing service during COVID-19 or have had no COVID-19 restrictions
22. Do most CR staff have the equipment (e.g., computer with video camera, remote access to hospital servers), space / privacy (e.g., home office with door) and other resources to be able to work efficiently from home during business hours?
    - Yes, they have everything they need
    - Yes, but only to do work that does not involve patients
    - No (if no, please specify: _________________________)
    - I do not know
23. Has there been a time when staff were concerned about delivering cardiac rehab due to lack of personal protective equipment?
    - Yes
    - No
    - I do not know
24. Has your program had any suspected or COVID-19 positive patients?
    - Yes
    - No
    - I do not know
25. How great do you perceive your risk has been to contact COVID-19 through the course of your work in CR? (enter a value from 0-100%) _____ %
26. Have you felt the need to work despite potential risk of contracting COVID-19 for fear of losing your job or need for pay?
    - Yes
    - No
27. Would your employer reduce your pay if you could not work due to having COVID-19 symptoms? (i.e, no paid sick leave)
    - Yes
    - No
28. Do you perceive your institution is giving you the COVID-19 related information you need to safely perform your duties and to ensure the safety of your patients?
    - Yes
    - No (if no, please specify: ________________________________)
29. Are you suffering personally due to COVID, in a way that is or could potentially impact your day-to-day work? (check all that apply)
    - Yes, I am feeling anxious at times
    - Yes, I am feeling isolated and lonely at times
    - Yes, I am feeling depressed or hopeless at times
    - Yes, I have been very worried about exposing my family to COVID-19
    - Yes, I have been really worried about some of my patients
    - Yes, I have found the workload to be really high to transition from delivering face-to-face services
    - Yes, I have been having trouble sleeping at night
    - Yes, I have kids and need to provide childcare during business hours or I need to take care of ill / older family members
    - Yes, I have had to or tried to stay away from my home for fear of spreading COVID
    - Yes, I am feeling _________ (other feelings – please specify)
    - No, I have been coping alright
    - I have been using this time to catch up on things and do some learning
    - Not applicable as we are not providing service during COVID-19 or have had no COVID-19 restrictions
30. Does your institution have a policy regarding the circumstances under which regular services can resume?
    - No
    - Yes

If yes, please describe _

- Not applicable as we are not providing service during COVID-19 or have had no COVID-19 restrictions

31. Which of the following factors do you think should form the basis for resumption of face-to-face CR services in the face of COVID-19? (check all that apply)
    - Cases in the community have peaked in a first wave
    - No new cases in the region for 28 days
    - No new cases in the region for 7 days
    - Hospital impact in the intensive care units has been stabilized, so no CR staff needs to be redeployed
    - Patients can be screened for COVID-19 symptoms before every in-person visit
    - Hospital impact in the intensive care units has been stabilized, so ill CR patients could get needed treatment
    - Sufficient testing capability is available across the region (health system and community)
    - Staff have sufficient personal protective equipment available
    - A treatment has been identified
    - A vaccine is available
    - Governments are mandating people to stay at home as much as possible
    - More knowledge is available on the cardiac impacts of COVID-19 and how they can be mitigated
    - Public transit has put measures into place to minimize transmission (e.g., distancing, cleaning)
    - Our institution has policies in place to protect patients (e.g., masks, sufficient space for physical distancing at our center)
    - Programs have capacity to deliver most services remotely, so relatively few patients would need to be seen at once at the centre
    - Programs can deliver any components on-site individually (not in a group)
    - Other (please specify: _________________________)
32. Are there any resources you have been using to help with care decisions or delivery during COVID-19? If yes, please share them below (with weblinks where available) (free text)
33. How can the International Council of Cardiovascular Prevention and Rehabilitation help you to adapt your services during COVID-19? (free text)

**Thank you most sincerely on behalf of the International Council of Cardiovascular Prevention and Rehabilitation for the time and expertise you have committed to complete this important questionnaire. We will post results and resources on our website soon:** <http://globalcardiacrehab.com/major-initiatives/covid-19-2/>
