## Appendix 2 for "Impacts of the COVID-19 Pandemic on Cardiac Rehabilitation Delivery around the World"

Appendix 2: Patient education mode and providers delivering, under usual practice and during COVID-19

| ***Who delivers the education*** | **Under usual practice** | **During COVID-19** | **Not Offered** |
| --- | --- | --- | --- |
| Nurses | 489 (73.1%) | 101 (15.1%) | 79 (11.8%) |
| Exercise specialist | 486 (75.0%) | 84 (13.0%) | 78 (12.0%) |
| Healthcare providers from multiple disciplines | 483 (75.8%) | 56 (8.8%) | 98 (15.4%) |
| Physician | 250 (43.0%) | 41 (7.0%) | 291 (50.0%) |
| ***Mode of Delivery*** | **Yes, and continue to offer during COVID-19** | **Yes, but not during COVID-19** | **No** |
| Telephone-based sessions (individual or group) | 385 (52.2%) | 66 (8.9%) | 287 (38.9%) |
| E-mailing resources as needed | 360 (49.1%) | 70 (9.5%) | 303 (41.3%) |
| Online resources for patients to view/download (asynchronous) | 352 (47.4%) | 89 (12.0%) | 302 (40.6%) |
| Mailing (postal service) resources as needed | 341 (47.1%) | 97 (13.4%) | 287 (39.5%) |
| One-on-one sessions | 280 (37.2%) | 356 (47.3%) | 117 (15.5%) |
| Virtual real-time/live interactive session (one-to-one) | 143 (19.6%) | 77 (10.6%) | 508 (69.8%) |
| Virtual real-time/live sessions (group) | 98 (13.4%) | 90 (12.3%) | 544 (74.3%) |
| Group sessions | 79 (10.7%) | 527 (71.5%) | 131 (17.8%) |
| Other | 28 (9.9%) | 13 (4.6%) | 243 (85.6%) |

Note: frequency and valid percentage shown.
